# Damaging missense variants in *IGF1R* implicate a role for IGF-1 resistance in the aetiology of type 2 diabetes

**DOI:** 10.1101/2022.03.26.22272972

**Authors:** Eugene J. Gardner, Katherine A. Kentistou, Stasa Stankovic, Samuel Lockhart, Eleanor Wheeler, Felix R. Day, Nicola D. Kerrison, Nicholas J. Wareham, Claudia Langenberg, Stephen O’Rahilly, Ken K. Ong, John R. B. Perry

## Abstract

Type 2 diabetes (T2D) is a chronic metabolic disorder with a significant genetic component. While large-scale population studies have identified hundreds of common genetic variants associated with T2D susceptibility, the role of rare (minor allele frequency < 0.1%) protein coding variation is less clear. To this end, we performed a gene burden analysis of 18,691 genes in 418,436 (n=32,374 T2D cases) individuals sequenced by the UK Biobank (UKBB) study to assess the impact of rare genetic variants on T2D risk. Our analysis identified T2D associations at exome-wide significance (P < 6.9×10^-7^) with rare, damaging variants within previously identified genes including *GCK, GIGYF1, HNF1A*, and *TNRC6B*. In addition, individuals with rare, damaging missense variants in the genes *ZEB2* (N=31 carriers; OR=5.5 [95% CI=2.5-12.0]; p=6.4×10^-7^), *MLXIPL* (N=245; OR=2.3 [1.6-3.2]; p=3.2×10^-7^), and *IGF1R* (N=394; OR=2.4 [1.8-3.2]; p=1.3×10^-10^) have higher risk of T2D. Carriers of damaging missense variants within *IGF1R* were also shorter (-2.2cm [-1.8-2.7]; p=1.2×10^-19^) and had higher circulating protein levels of insulin-like growth factor-1 (IGF-1; 2.3 nmol/L [1.7-2.9] p=2.8×10^-14^), indicating relative IGF-1 resistance. A likely causal role of IGF-1 resistance on T2D was further supported by Mendelian randomisation analyses using common variants. Our results increase our understanding of the genetic architecture of T2D and highlight a potential therapeutic benefit of targeting the Growth Hormone/IGF-1 axis.

## Introduction

Type 2 diabetes (T2D) is a complex disease characterised by insulin resistance and beta-cell dysfunction. An estimated 630 million adults are expected to have T2D by 2045^1^ making it one of the fastest growing global health challenges of the 21st century. Genome-wide association studies (GWAS) have successfully identified more than 500 genomic loci to be associated with T2D^2^, although the majority of these are driven by common variants with small individual effects on T2D risk.

Over 90% of GWAS loci lie in non-coding regions of the genome^3^, presenting a major hurdle for the identification of the underlying causal genes and the translation of these findings into mechanistic insight. In contrast, analysis of rare protein-coding variation captured by DNA sequencing has the potential to more directly implicate individual genes and biological mechanisms. The UK Biobank (UKBB)^4^ study recently made Exome Sequencing (ES) data available for 454,787 UKBB participants^5^. This offers an unprecedented opportunity to explore the contribution of rare coding variation to the risk of T2D with much greater power than previously possible^6–8^. Initial exome-wide association analyses of these data have identified gene-based associations with increased risk of T2D for *GCK, HNF1A, HNF4A, GIGYF1, CCAR2, TNRC6B* and *PAM*, and protective effects for variants in *FAM234A* and *MAP3K15*^*5,9–14*^.

In this study, we combined multiple sources of health record data to identify additional T2D cases and used an extended range of variant classes and allele frequency cutoffs in order to directly implicate novel genes in the aetiology of T2D. Our results highlight a number of previously missed associations and support a role for Insulin-like Growth Factor 1 (IGF-1) resistance in the pathogenesis of T2D.

## Results

### Exome-wide burden testing in the UK Biobank

To identify genes associated with T2D risk, we performed an Exome-wide association study (ExWAS) using ES data derived from 418,436 European genetic-ancestry UKBB participants^5^. As our primary outcome, we identified 32,374 (7.7%) participants with likely incident or prevalent T2D using phenotype curation that integrated multiple data sources, including hospital episode statistics, self-reported conditions, death records, and use of T2D medication (see methods).

Individual gene burden tests were performed by collapsing genetic variants across 18,691 protein-coding genes in the human genome. We tested four functional categories across two population prevalences (minor allele frequency < 0.1% and singletons), including high-confidence Protein Truncating Variants (PTVs), missense variants stratified by two REVEL score thresholds^15^, and synonymous variants as a negative control (Figure 1; methods). We identified 13 gene-functional annotation pairs with 30 or more rare allele carriers, representing 7 non-redundant genes, associated with T2D at exome-wide statistical significance (p < 6.9×10^-7^; Supplementary Table 1; methods). Our results are statistically well-calibrated, both as indicated by low exome-wide inflation scores (e.g. PTV λ=1.047) and by the absence of significant associations with synonymous variant burden (Figure 1B-E; Supplementary Figure 1). To ensure our results were not biassed by our approach, we implemented burden tests using STAAR^16^ and a logistic model and arrived at substantially similar conclusions (Supplementary Figure 2; Supplementary Table 1; methods).

**Figure 1.**
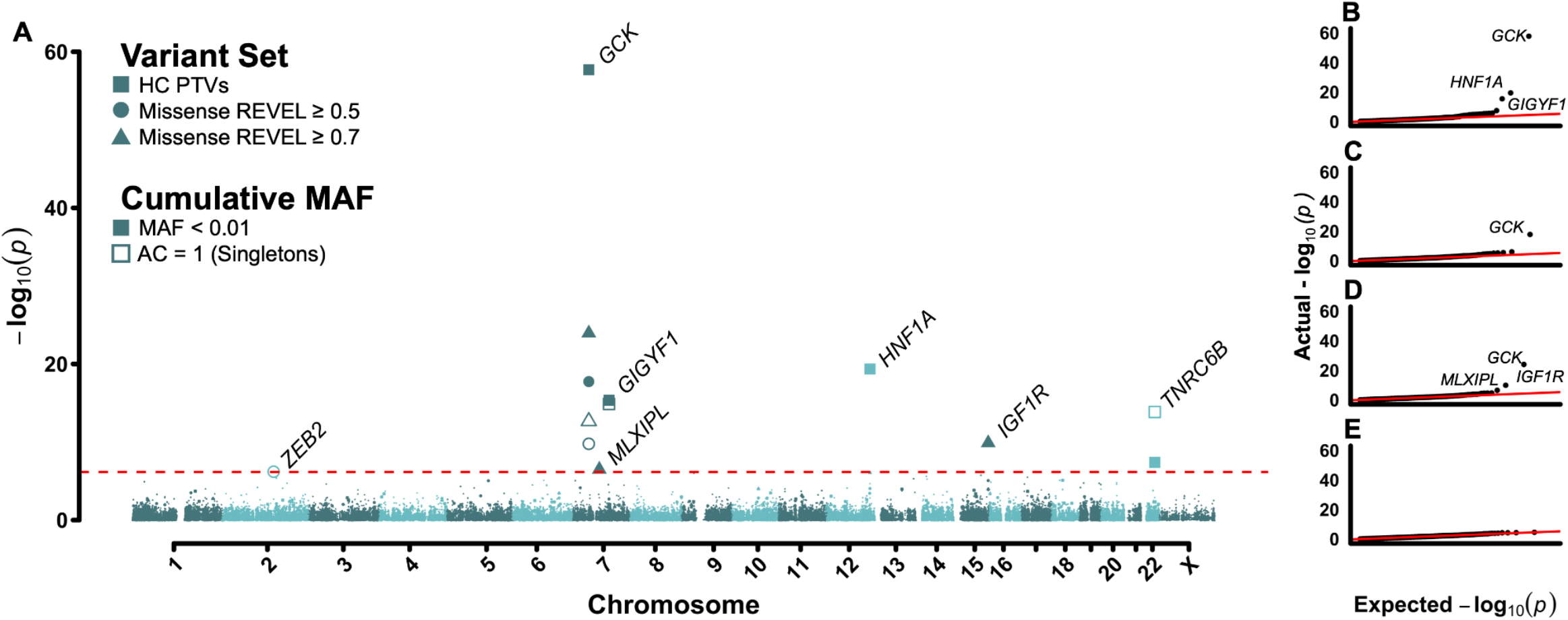
Exome-wide association results for T2D. (A) Manhattan plot displaying results of gene burden tests for T2D risk. Genes passing exome-wide significance (p < 6.9×10^-7^) are labelled. Point shape indicates variant class tested. (B-E) QQ plots for (B) high confidence PTVs (C) REVEL ≥ 0.5 Missense Variants (D) REVEL ≥ 0.7 missense variants and (E) synonymous variants (negative control).

We confirmed the T2D associations at all three genes identified by three previous studies of T2D risk that incorporated European genetic-ancestry individuals from the UKBB study^11,12,14^: *GCK* (N=35 carriers; OR=58.5 [95% CI=25.5-134.5]; p=2.0×10^-58^), *HNF1A* (N=33; OR=12.7 [6.2-25.8]; p=4.4×10^-20^), and *GIGYF1* (N=133; OR=4.7 [3.1-7.0]; p=4.4×10^-16^; Figure 2). As in these previous studies, we similarly found that carriers of PTVs within these genes had substantially increased risk for developing T2D (Figure 1A).

**Figure 2.**
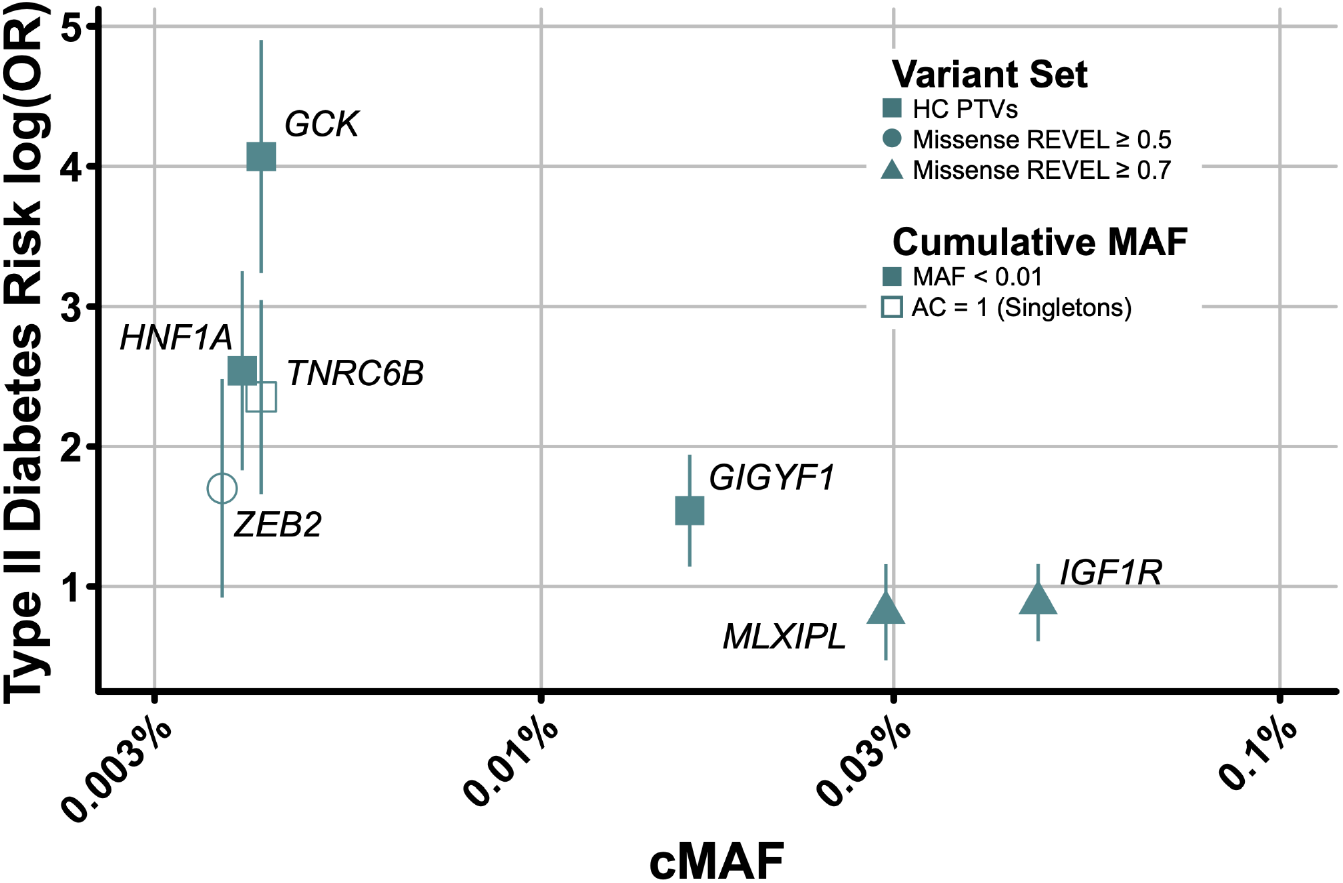
Relationship between cumulative minor allele frequency and odds ratio for T2D. Plotted is T2D risk as quantified by log(Odds Ratio) versus cumulative minor allele frequencies (cMAF) for genes significantly associated with T2D risk. For each gene, only the most significantly associated variant mask is shown. Error bars indicate 95% confidence intervals.

We also confirmed the T2D association at *TNRC6B* (N=35; OR=10.5 [5.3-21.0]; p=1.4×10^-14^), which was previously reported as ‘potentially spuriously associated’ with T2D risk^11^; several new lines of evidence provide confidence in this association. Firstly, our result is not attributable to a single variant of large effect as evidenced by the strength of association with singleton variants (Figure 1A). Secondly, aside from a single individual carrying two balanced deletions, inspection of the underlying ES reads did not reveal a markedly increased error rate in variant calling or genotyping in *TNRC6B* as was suggested by Deaton et al.^11^. Thirdly, the association persisted after excluding 14 individuals who carry a singleton PTV in a potentially non-constitutive exon as measured by PEXT (p=3.6×10^-7^)^17^. Finally, we also found that *TNRC6B* PTV carriers had elevated HbA1c levels when considering both T2D cases (4.1 mmol/mol [2.5-5.7]; p=7.2×10^-7^; Supplementary Figure 3) and controls (1.6 mmol/mol [0.2-2.1]; p=1.8×10^-2^), consistent with the elevated long-term blood glucose levels observed in T2D patients.

We also identified three additional genes that, when disrupted by rare genetic variation (minor allele frequency < 0.1% or singletons), are associated with increased T2D risk: *IGF1R* (N=394; OR=2.4 [1.8-3.2]; p=1.3×10^-10^), *MLXIPL* (N=245; OR=2.3 [1.6-3.2]; p=3.2×10^-7^), and *ZEB2* (N=31; OR=5.5 [2.5-12.0]; p=6.4×10^-7^; Figure 1). Unlike previously reported genes outlined above, damaging missense variants but not PTVs in these genes were associated with T2D risk (Figure 2). Indeed, at these genes T2D associations were apparent only with missense variants with high REVEL scores (≥0.7), or those variants considered to be the most damaging as per current (2020) Association for Clinical Genomic Science guidelines.

Specifically, and as expected, we found that carriers of PTVs within *GCK, GIGYF1*, and *HNF1A* all had significantly elevated circulating glucose and HbA1c levels. Among novel genes, *IGF1R* missense carriers had nominally higher HbA1c levels (1.1 mmol/mol [0.6-1.6]; p=3.7×10^-6^).

### Exploring Common Variant Associations at Highlighted Genes

We next attempted to cross-validate the rare-variant associations for all seven exome-wide significant genes by identifying proximal common variants (±50kb of a gene’s coding sequence) previously reported to be associated with related glycaemic or metabolic phenotypes (methods; Supplementary Table 2). Four genes fell within glycaemic trait associated loci and all seven overlapped known metabolic trait associations. For several of these common variant-phenotype combinations, we also identified an association with rare variant burden (Supplementary Figure 3). Additionally, three of the four novel genes we report here were identified in the most recent publicly available T2D GWAS^2^ as being either the closest or most likely causal gene for a common variant genome-wide significant signal: *IGF1R, TNRC6B*, and *ZEB2* (Supplementary Table 2).

Notably, common non-coding variants at the *IGF1R* locus have been previously reported for T2D and fasting glucose^2,18^. The lead fasting glucose-associated SNP (rs6598541-A; p= 4×10^-12^) was associated with 0.0114 mmol/L [0.0097-0.0131] higher levels of glucose, while the lead T2D SNP (rs59646751-T; p=4×10^-9^) increases risk of T2D by an odds ratio of 1.024 [1.020-1.028]. Both SNPs are intronic in *IGF1R*, in moderate LD in European populations (R^2^=75.5%)^19^, and are eQTLs for *IGF1R*^*20*^. Furthermore, at both SNPs the *IGF1R* expression-lowering alleles are associated with higher levels of circulating IGF-1 (p=7×10^-7^ and 9×10^-7^, respectively; Figure 3)^21^ and with higher T2D risk and fasting glucose.

**Figure 3.**
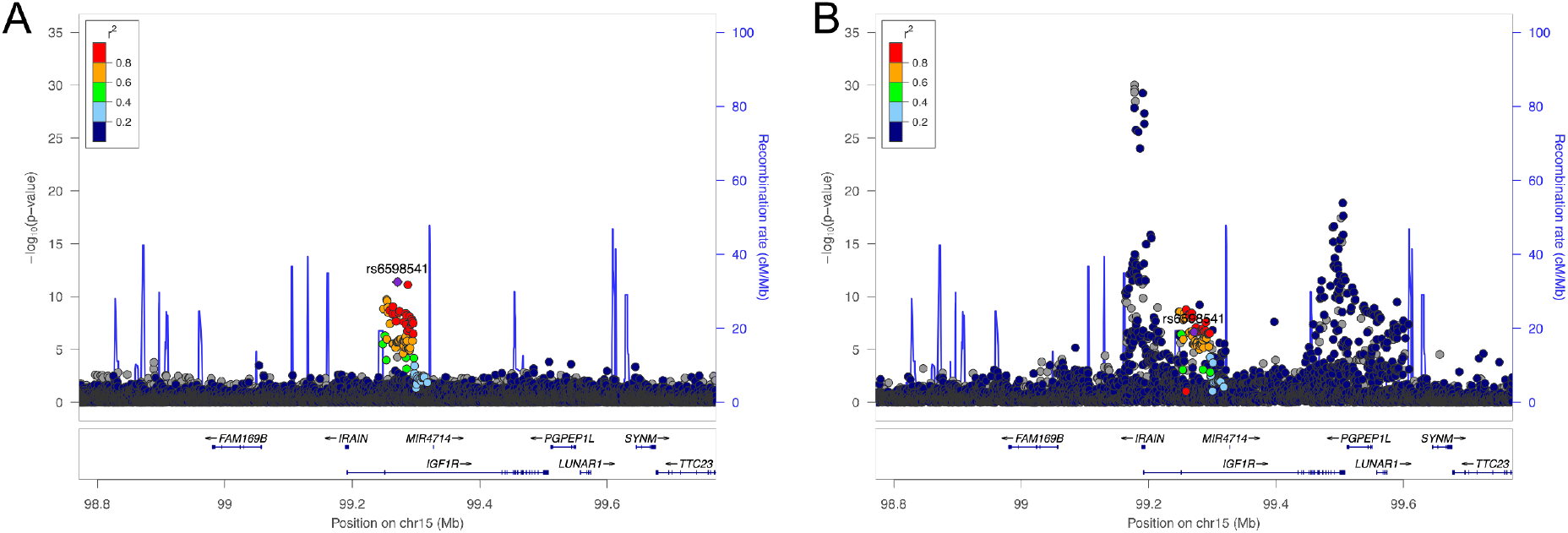
Common variant associations at the IGF1R locus. Association pattern between SNPs at the IGF1R locus and Fasting Glucose levels (A) and IGF-1 levels (B).

### Interrogating *IGF1R* and Risk for T2D

To understand how rare damaging missense variants in *IGF1R* lead to increased risk of T2D, we performed burden tests for circulating IGF-1 levels and anthropometric traits. We found that carriers of damaging missense variants in *IGF1R* had increased circulating IGF-1 levels (2.1 nmol/L [1.5-2.6]; p=1.9×10^-14^), but shorter adult stature (-2.2cm [-1.8-2.7]; p=1.2×10^-19^), and lower relative height at age 10 (p=1.1×10^-7^). These findings indicate that carriers of rare damaging missense variants in *IGF1R* that increase risk of T2D have relative IGF-1 resistance.

To explore how damaging missense variants disrupt *IGF1R* function, we next categorised variants by protein domain. Carriers of qualifying variants within the *IGF1R* protein kinase (residues 999-1274)^22^ had a higher risk for T2D (N= 179; OR=3.4 [2.3-4.9]; p=1.9×10^-10^) than those with qualifying variants outside this domain (N=215; OR=1.7 [1.2-2.6]; p=8.2×10^-3^). We thus conclude that dysfunction within the protein kinase domain could decrease downstream signal transduction resulting in IGF-1 resistance. This may also explain why, despite the relatively large number of *IGF1R* (N=64) PTV carriers in the UKBB, we did not find that *IGF1R* PTV carriers had increased T2D risk. When bound by IGF-1 and to induce downstream signal transduction, *IGF1R* functions as a homo or heterodimer (i.e. with *INSR* as a hybrid receptor)^23^. As half of a missense carrier’s *IGF1R* molecules will contain errors in the protein kinase domain, dimerisation will incorporate at least one defective molecule 75% of the time and therefore lead to reduced downstream signal transduction. In the case of PTV carriers, since one copy is likely missing due to nonsense-mediated decay, dimerisation will always incorporate two functional copies. Therefore, the association of damaging *IGF1R* missense variants with T2D may be due to a dominant-negative effect rather than decreased protein abundance; however, additional functional studies are ultimately required to confirm the mechanism underlying these variants.

To explore whether rare variants in other components of the GH-IGF1 hormone pathway might influence T2D risk, we next identified a further nine genes in the GH-IGF1 pathway that showed gene-burden associations with circulating IGF-1 levels in any of our burden tests (Supplementary Table 3), including seven genes with known roles in regulating GH secretion or GH signalling and three genes with known roles in IGF-1 bioavailability. We tested their associations with childhood and adult height to indicate the functional relevance of rare variation in these genes. None of the seven GH-related genes showed any association with T2D. Rare damaging variants in *IGFALS*, which encodes a component of the IGF-1 ternary complex, lowered lower circulating IGF-1 and were nominally associated with shorter childhood height (indicative of lower IGF-1 bioactivity) and higher risk of T2D. Rare damaging variants in *IGFBP3* (the major IGF binding protein), which lowered circulating IGF-1 levels, were nominally associated with taller childhood height (indicative of higher IGF-1 bioactivity) and lower risk of T2D. Hence damaging rare variants that disrupt IGF-1 bioactivity, but not those that alter GH secretion or signalling, appear to increase T2D risk.

### Causality of IGF-1 Levels with T2D Risk

A previous epidemiological study described a protective association between baseline circulating IGF-1 protein levels and incident T2D^24^. However, subsequent similar studies found no such association^25,26^ and conversely a previous study that modelled common genetic variants in a Mendelian randomization framework inferred an adverse causal effect of higher circulating IGF-1 levels on T2D^27^.

To explore this apparent inconsistency, we examined the likely causal role of IGF-1 on T2D by modelling 784 independent genetic signals for circulating IGF-1 levels identified in 428,525 white European UKBB individuals^21^ and summary statistics from the largest reported GWAS meta-analysis of T2D^28^. We confirmed the previously reported^27^ association between genetically-predicted higher IGF-1 levels and higher risk of T2D in inverse-variance weighted (IVW; OR=1.105 per SD [95% CI 1.039-1.170]; p=2.9×10^-3^) and sensitivity models (Supplementary Table 4). However, we noted substantial heterogeneity in the relationships between individual IGF-1 signals and T2D (I-square=85.7%) as well as in their associations with adult height (IVW Beta=0.142; p=8.9×10^-9^; I-square=97.7%). Among the common genetic instruments for higher circulating IGF-1 levels, individual variants at the *IGF1* locus (rs11111274) and the *IGF1R* locus (rs1815009) show directionally-opposite effects on childhood height and T2D (taller height and lower T2D risk for IGF1; shorter height and higher T2D risk for *IGF1R*; Supplementary Figure 4). Hence, reported common variant instruments for higher IGF-1 levels comprise a mixture of functionally-opposing signals, i.e. higher levels of bioactive IGF-1 but also higher IGF-1 resistance.

## Discussion

Here we present the results of an ExWAS to assess the contribution of rare variant burden to T2D risk (Figure 1). We identified three genes previously reported by a recent analysis of the UKBB (*GCK, HNF1A*, and *GIGYF1*)^14^, provide stronger evidence for a previously nominally associated gene (*TNRC6B*)^11^, and identified three new genes (*ZEB2, MLXIPL*, and *IGF1R*) where rare variants increase susceptibility to T2D (Figure 2). Using publicly available data, we showed that common variation nearby these genes is associated with a wide range of glycemic and metabolic traits and (Figure 3; Supplementary Table 2)^2,18^, providing further support for these rare variant associations. We further interrogated rare and common variant associations to show that disruption of *IGF1R* due to damaging missense variants in the cytoplasmic protein kinase domain leads to IGF-1 resistance and higher T2D risk. Overall, our results implicate a wider protective effect of IGF-1 bioactivity on susceptibility to T2D.

While our results are complementary to previous ExWAS^11,14^, we clarified evidence linking *TNRC6B* to T2D and identified three additional genes missed by previous analyses of the UKBB. A key advantage of our approach was to carefully curate multiple data sources to identify and validate T2D cases. Furthermore, we used a different genetic analytical approach to those previous studies. Nag et al.^14^ limited their burden testing to either PTVs, the findings of which we replicate here, or to missense variants with comparatively low deleteriousness scores (REVEL > 0.25 or Missense Tolerance Ratio intragenic percentiles ≤ 50%). In this study, we have shown the benefit of considering missense variants computationally predicted to be severely damaging (REVEL ≥ 0.5 and 0.7)^15^. While such variants are much rarer in the population – only ∼8% of missense variants in UKBB have REVEL scores ≥ 0.7 – they are much more likely to disrupt protein function and thus increase risk for disease. These conclusions are similar to those shown previously for anthropometric traits^10^, which have shown a relationship between PTVs in *IGF1R* and several growth measures, but not for damaging missense variants.

A key finding of our work is the novel association between *IGF1R* and T2D risk. Loss of function mutations in *IGF1R* have been reported in children presenting with intra-uterine growth restriction, short stature and elevated IGF-1 levels^29–31^. Our findings of rare damaging variants at *IGF1R*, and also at *IGFALS* and *IGFBP3*, indicate that reduced IGF-1 bioactivity and signalling increases risk for T2D. There are several plausible mechanisms to link *IGF1R* to T2D. *IGF1R*, responding to both systemic and locally generated IGF-1, may play a role in the development of several tissues central to the control of glucose metabolism including pancreatic islets, adipose tissue and skeletal muscle^32^. An alternative explanation involves the complex relationship between growth hormone (GH) and IGF-1. GH, produced in a highly controlled and pulsatile manner from the somatotropes of the anterior pituitary, is the major stimulus to the hepatic expression and secretion of IGF-1, the major source of this circulating hormone. GH also has metabolic effects that are independent of IGF-1, largely exerted by its powerful lipolytic effects in adipose tissue^33–38^, which if uncontrolled can lead to the accumulation of ectopic lipid in non-adipose tissue resulting in insulin resistance. This is elegantly demonstrated by studies in mice in which IGF-1 is selectively deleted in the liver^39,40^. These mice show a striking increase in circulating GH levels, accompanied by marked insulin resistance which is entirely abrogated by the blockade of GH signalling. This model can explain the insulin resistance and frequent T2D seen in conditions such as acromegaly, where GH and IGF-1 levels are persistently raised due to a functional somatotrope tumour^41^, and the striking protection from T2D seen in patients with Laron Dwarfism, whose markedly reduced circulating IGF-1 levels are due to biallelic LoF mutations in the GH receptor^42^. Loss of function mutations in IGF1R are likely to result in compensatory increases in GH secretion, and consequently higher levels of circulating IGF-1 that we observed in the carriers of such mutations. While this may partially compensate for impairment in IGF1R function, the IGF1R-independent effects of GH are likely to have a deleterious effect on systemic glucose metabolism. Of note in this regard, a single human proband with a homozygous loss of function mutation in IGF-1 had elevated circulating GH and severe insulin resistance^43,44^. Therapy with exogenous IGF-1 resulted in suppression of GH and a dose dependent improvement in insulin sensitivity^44^. Accordingly, genetically reduced GH secretion and signalling would lead to reduced IGF-1 bioactivity, but without the consequent effects of elevated GH on fatty acid metabolism and insulin resistance, and hence no alteration in T2D risk. We propose that currently available drugs which reduce GH secretion or block its action may have metabolic benefits in patients with T2D and damaging missense variants in the protein kinase domain of *IGF1R*.

Our findings also demonstrate the challenge of interpreting Mendelian randomisation results of circulating biomarkers. Elevated levels may reflect higher levels of secretion and biomarker activity, but are also increased by mechanisms that reduce biomarker bioavailability or sensitivity. Hence, genetic instruments for higher biomarker levels may comprise a mixture of markers for both higher and lower biomarker activity. To distinguish these actions, we suggest that individual common variants are first tested for association with some indicator of biomarker activity (i.e. childhood height as an indicator of IGF-1 activity).

Our rare variant analysis also implicates *MLXIPL* as a T2D susceptibility gene for the first time. *MLXIPL* encodes the carbohydrate response element binding protein (CHREBP), a transcription factor that acts in concert with its obligate binding partner MLX to regulate the cellular response to carbohydrate^45–47^ and is highly expressed in liver, fat, and muscle. Global or tissue-specific ablation of *MLXIPL* in mice impairs insulin sensitivity^48–51^. Common variants at the *MLXIPL* locus associate with *SHBG*, a biomarker of insulin sensitivity^52^ and with serum triglycerides^53,54^. Notably, *MLXIPL* is one of the 26-28 genes deleted in Williams Syndrome, the result of a deletion of contiguous genes on chromosome 7q11.23. Patients with this syndrome are characterised by marked insulin resistance and an increased risk of diabetes^55^. It seems likely that haploinsufficiency for *MXLIPL* contributes significantly to the metabolic disturbances characteristic of Williams syndrome.

Overall, our findings suggest that deeper interrogation of multiple variant types when performing ExWAS can and will lead to the discovery of additional genes associated with a wide-range of human diseases.

## Methods

### UK Biobank Data Processing and Quality Control

To conduct rare variant burden analyses outlined in this publication, we queried ES data for 454,787 individuals provided by the UKBB study^5^. Individuals were excluded from further analysis if they had excess heterozygosity, autosomal variant missingness on genotyping arrays ≥ 5%, or were not included in the subset of phased samples as defined in Bycroft et al.^56^. We further excluded all study participants who were not of broadly European genetic ancestry, leaving a total of 421,065 individuals for further analysis.

To perform variant quality control and annotation, we utilised the UKBB Research Analysis Platform (RAP; https://ukbiobank.dnanexus.com/). The RAP is a cloud-based compute environment which provides a central data repository for UKBB ES and phenotypic data. Using bespoke applets designed for the RAP, we performed additional quality control of ES data beyond that already documented in Backman et al.^5^. Using provided population-level Variant Call Format (VCF) files, we first split and left-corrected multi-allelic variants into separate alleles using ‘bcftools norm’^57^. Next, we performed genotype-level filtering using ‘bcftools filter’ separately for Single Nucleotide Variants (SNVs) and Insertions/Deletions (InDels) using a missingness-based approach. With this approach, SNV genotypes with depth < 7 and genotype quality < 20 or InDel genotypes with a depth < 10 and genotype quality < 20 were set to missing (i.e. ./.). We further tested for an expected alternate allele contribution of 50% for heterozygous SNVs using a binomial test; SNV genotypes with a binomial test p. value ≤ 1×10^-3^ were set to missing. Following genotype-level filtering we recalculated the proportion of individuals with a missing genotype for each variant and filtered all variants with a missingness value > 50%.

We next annotated variants using the ENSEMBL Variant Effect Predictor (VEP) v104^58^ with the ‘--everything’ flag and plugins for REVEL^15^, CADD^59^, and LOFTEE^60^ enabled. For each variant, we prioritised a single ENSEMBL transcript based on whether or not the annotated transcript was protein-coding, MANE select v0.97, or the VEP Canonical transcript, respectively. Individual consequence for each variant was prioritised based on severity as defined by VEP. Following annotation, we grouped stop gained, frameshift, splice acceptor, and splice donor variants into a single Protein Truncating Variant (PTV) category. Missense and synonymous variant consequences are identical to those defined by VEP. Only autosomal or chrX variants within ENSEMBL protein-coding transcripts and within transcripts included on the UKBB ES assay were retained for subsequent burden testing.

### Exome-wide association analyses in the UK Biobank

To perform rare variant burden tests using filtered and annotated ES data, we employed a custom implementation of BOLT-LMM v2.3.6^61^ for the RAP. BOLT-LMM expects two primary inputs: i) a set of genotypes with minor allele count > 100 derived from genotyping arrays to construct a null model and ii) a larger set of imputed variants to perform association tests. For the former, we queried genotyping data available on the RAP and restricted to an identical set of individuals used for rare variant association tests. For the latter, and as BOLT-LMM expects imputed genotyping data as input rather than per-gene carrier status, we created dummy genotype files where each variant represents one gene and individuals with a qualifying variant within that gene are coded as heterozygous, regardless of the number of variants that individual has in that gene. To test a range of variant annotation categories across the allele frequency spectrum, we created dummy genotype files for minor allele frequency < 0.1% and singleton high confidence PTVs as defined by LOFTEE, missense variants with REVEL ≥ 0.5, missense variants with REVEL ≥ 0.7, and synonymous variants. For each phenotype tested, BOLT-LMM was then run with default parameters other than the inclusion of the ‘lmmInfOnly’ flag. When exploring the role of rare variants in the IGF-1/GH axis and to incorporate less deleterious missense variants, we also used an additional set of variant annotations which combined missense variants with CADD ≥ 25 and high confidence PTVs (i.e. Damaging; Supplementary Table 3). To derive association statistics for individual variants, we also provided all 26,657,229 individual markers regardless of filtering status as input to BOLT-LMM. All tested phenotypes were run as continuous traits corrected by age, age^2^, sex, the first ten genetic principal components as calculated in Bycroft et al.^56^, and study participant ES batch as a categorical covariate (either 50k, 200k, or 450k). For phenotype definitions used in this study, please refer to Supplementary Table 5. Only the first instance (initial visit) was used for generating all phenotype definitions unless specifically noted in Supplementary Table 5.

To provide an orthogonal approach to validate our BOLT-LMM results, we also performed per-gene burden tests with STAAR^16^ and a generalised linear model as implemented in the python package ‘statsmodels’^62^. To run STAAR, we created a custom Python and R workflow on the RAP. VCF files were first converted into a sparse matrix suitable for use with the R package ‘Matrix’ using ‘bcftools query’. Using the ‘STAAR’ R package, we first ran a null model with identical coefficients to BOLT-LMM and a sparse relatedness matrix with a relatedness coefficient cutoff of 0.125 as described by Bycroft et al.^56^. We next used the function ‘STAAR’ to test all protein-coding transcripts as outlined above. To run generalised linear models, we used a three step process. First, we ran a null model with all dependent variables as continuous traits, corrected for control covariates identical to those included in BOLT-LMM. Next, using the residuals of this null model, we performed initial regressions on carrier status to obtain a preliminary p. value. Finally, for individual genes that passed a lenient p. value threshold of <1×10^-4^, we recalculated a full model to obtain exact test statistics with family set to ‘binomial’ or ‘gaussian’ if the trait was binary or continuous, respectively. Generalised linear models utilised identical input to BOLT-LMM converted to a sparse matrix.

### Common variant GWAS lookups

Common variant associations at the identified genes were queried using the T2D Knowledge Portal (https://t2d.hugeamp.org) and the Open Targets Genetics platform (https://genetics.opentargets.org/)^63^. Trait associations from the T2D Knowledge Portal are presented in Supplementary Table 2 and were only included if the paired gene was assigned as the nearest gene to the association signal as a crude proxy for causality. Accompanying HuGE scores were extracted for the highest-scoring glycaemic common variants associations. Locus2Gene scores based on data from Vujkovic et al.^2^ were extracted from the Open Targets Genetics platform and are presented in Supplementary Table 2. For the *IGF1R* locus follow-up, we used sentinel SNP information for Vujkovic et al.^2^ and summary statistics from the recent fasting glucose MAGIC meta-analysis^18^ and circulating IGF-1 levels GWAS. eQTL data was accessed through GTEx v8^20^. Effect estimates in the text have been aligned towards the T2D/glucose increasing alleles, using LD information from LDlink^19^. Regions in Figure 3 were plotted using LocusZoom^64^.

### Mendelian Randomisation Using IGF-1 levels

To examine the likelihood of a causal effect of IGF-1 on the risk of T2D, we applied Mendelian randomization (MR) analysis. In this approach, genetic variants that are significantly associated with an exposure of interest are used as instrumental variables (IVs) to test the causality of that exposure on the outcome of interest. For a genetic variant to be a reliable instrument, the following assumptions should be met: (1) the genetic instrument is associated with the exposure of interest, (2) the genetic instrument should not be associated with any other competing risk factor that is a confounder, and (3) the genetic instrument should not be associated with the outcome, except via the causal pathway that includes the exposure of interest^65^. As IVs, we used the 831 IGF-1 genome-wide significant signals reported in a recent GWAS on IGF-1^21^. As our outcome data, we selected the largest publicly available independent T2D dataset available in 893,130 European genetic ancestry individuals (9% cases) from Mahajan et al.^28^ If a signal was not present in the outcome GWAS, we searched the UKBB white European dataset for proxies (within 1 Mb and r^2^ > 0.5) and chose the variant with the highest r^2^ value, which left 784 independent markers for MR analysis. Genotypes at all variants were aligned to designate the IGF-1-increasing alleles as the effect alleles.

To conduct our MR analysis, we used the inverse-variance weighted (IVW) model as the primary model as it offers the most statistical power^66^; however, as it does not correct for heterogeneity in outcome risk estimates between individual variants^67^, we applied a number of sensitivity MR methods that better account for heterogeneity^68^. These include an Egger analysis to identify and correct for unbalanced heterogeneity (‘horizontal pleiotropy’), indicated by a significant Egger intercept (p<0.05)^69^, and weighted median (WM) and penalised weighted median (PWM) models to correct for balanced heterogeneity^70^. In addition, we introduced the radial method to exclude variants from each model in cases where they are recognized as outliers, as well as Steiger filtering to assess for potential reverse causality (i.e. variants with stronger association with the outcome than with the exposure)^71^. As previous work on IGF-1 showed a strong association with height, and to a lesser extent BMI^27^, we also used multivariable MR analysis^72^ to estimate the direct effect of IGF-1 levels on T2D not mediated by BMI or height by adjusting for their effects as covariates using queried phenotype data for UKBB participants (Supplementary Table 4; Supplementary Table 5). In order to examine the individual level effect of IGF1 and IGF1R loci on T2D, BMI, childhood and adult height, we performed the variant-specific lookups as well as calculated the Wald ratio using the R package ‘TwoSampleMR’^73^. All results presented in the main text are expressed in standard deviations of IGF-1 levels (one S.D. is equivalent to ∼5.5 nmol/L in UKBB). Values available in Supplementary Table 4 are raw data, per unit IGF-1.

## Data Availability

Data will be deposited to the UKBiobank returns catalogue on publication.

## Acknowledgements

This work was funded by the Medical Research Council (Unit programs: MC_UU_12015/2, MC_UU_00006/2, MC_UU_12015/1, and MC_UU_00006/1). S.L. is supported by a Wellcome Trust Clinical PhD Fellowship (225479/Z/22/Z). This research was supported by the NIHR Cambridge Biomedical Research Centre (BRC-1215-20014). For the purpose of open access, the author has applied a Creative Commons Attribution (CC BY) licence to any Author Accepted Manuscript version arising. This research was conducted using the UK Biobank Resource under application 9905.

