## Supplementary Materials for "Damaging missense variants in *IGF1R* implicate a role for IGF-1 resistance in the aetiology of type 2 diabetes"

### Supplementary Figures

#### Supplementary Figure 1

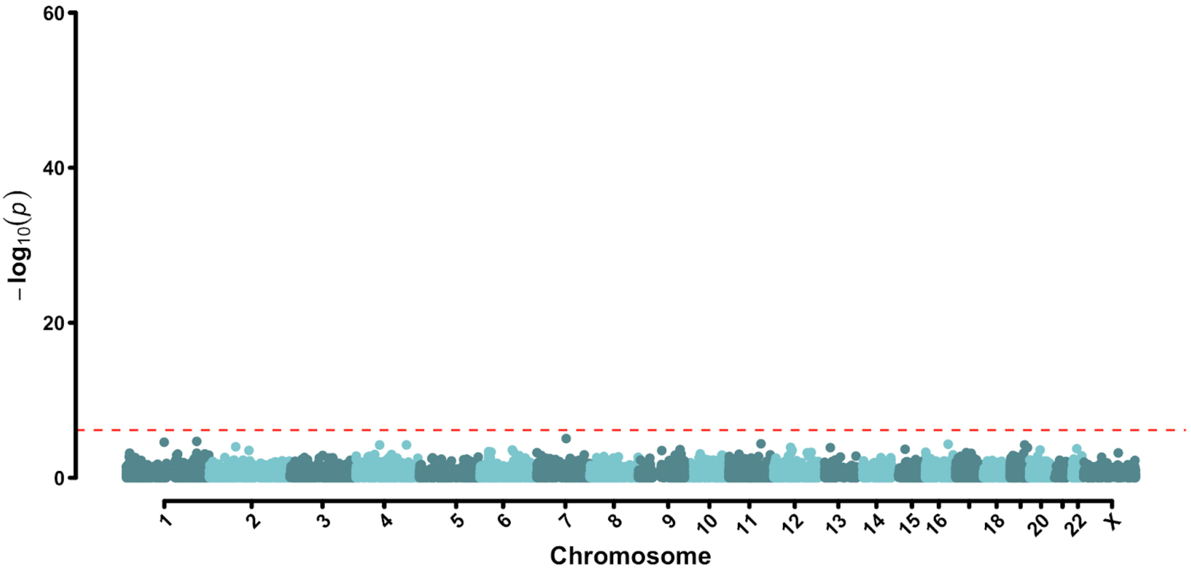


**Exome-wide Association Results for Synonymous Variants.** Plotted are per-gene burden results when only considering synonymous variants. The red line indicates our exome-wide significant p value after Bonferroni correction of $6.9 \times 10^{-7}$.

Supplementary Figure 2

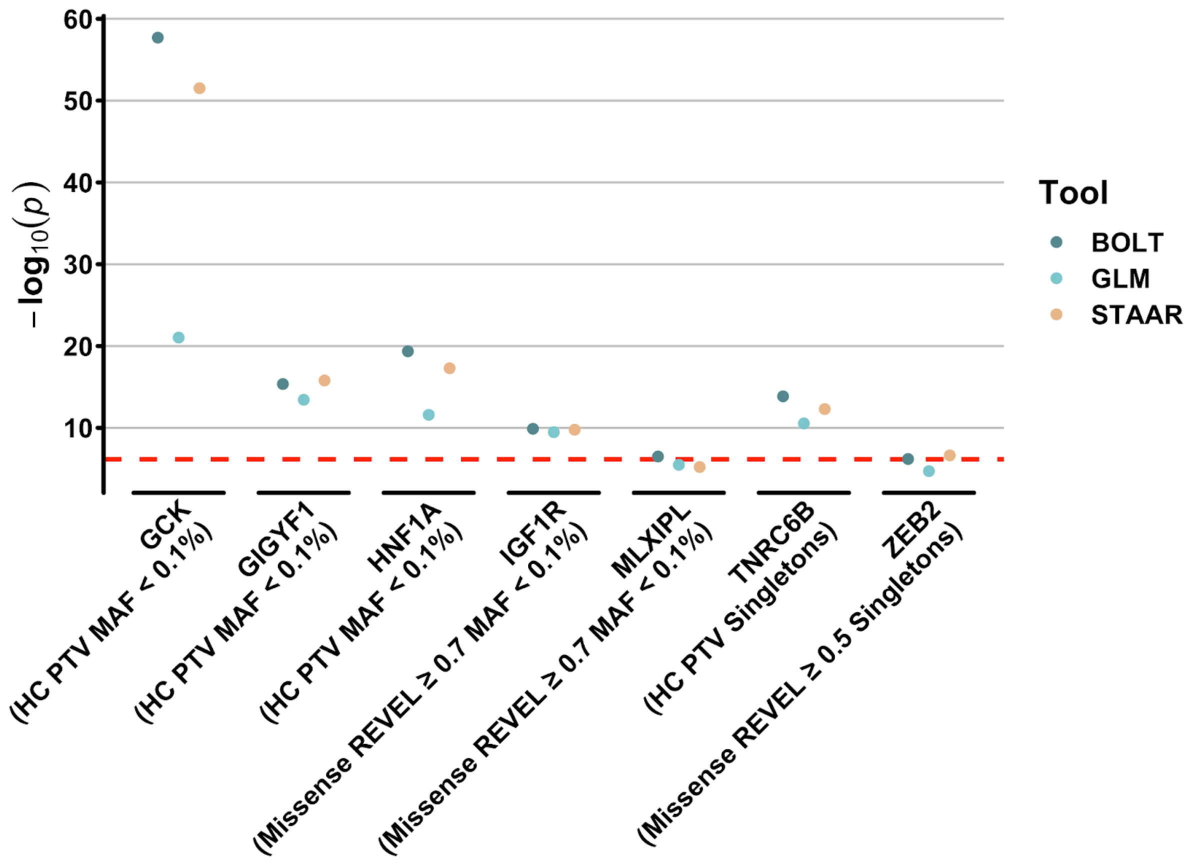


**Exome-wide association results for additional methods.** Displayed are association results for both our primary approach (BOLT) as well as two other orthogonal approaches (generalised linear models (GLM) and STAAR). -$\log_{10}$(p values) for each tool are plotted only for all genes identified as exome-wide significant with BOLT.

Supplementary Figure 3

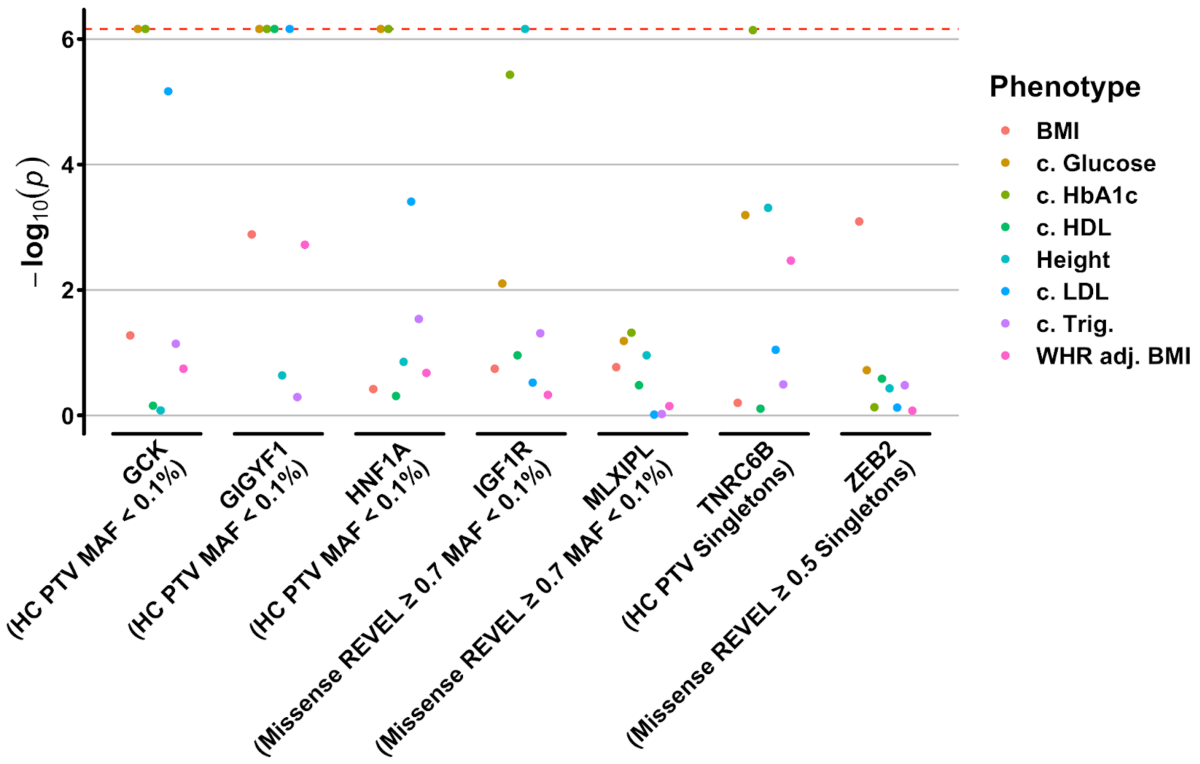


**Additional phenotypes associated with T2D or the GH-IGF1 pathway.** Shown are associations between rare variant burden and eight additional phenotypes with known associations to type 2 diabetes or the GH-IGF1 signalling pathway. Plotted are -$\log_{10}$ p. values for each trait derived from the mask-MAF cutoff most significantly associated with T2D (Main Text Figure 1; Supplementary Table 1). -$\log_{10}$ p. values are capped at exome wide significance (-$\log_{10}$ p = 6.16) to enable comparison of nominally associated signals. ‘c.’, ‘trig.’, and ‘adj.’ in the figure legend stand for ‘circulating’, ‘triglycerides’, and ‘adjusted’, respectively.

Supplementary Figure 4

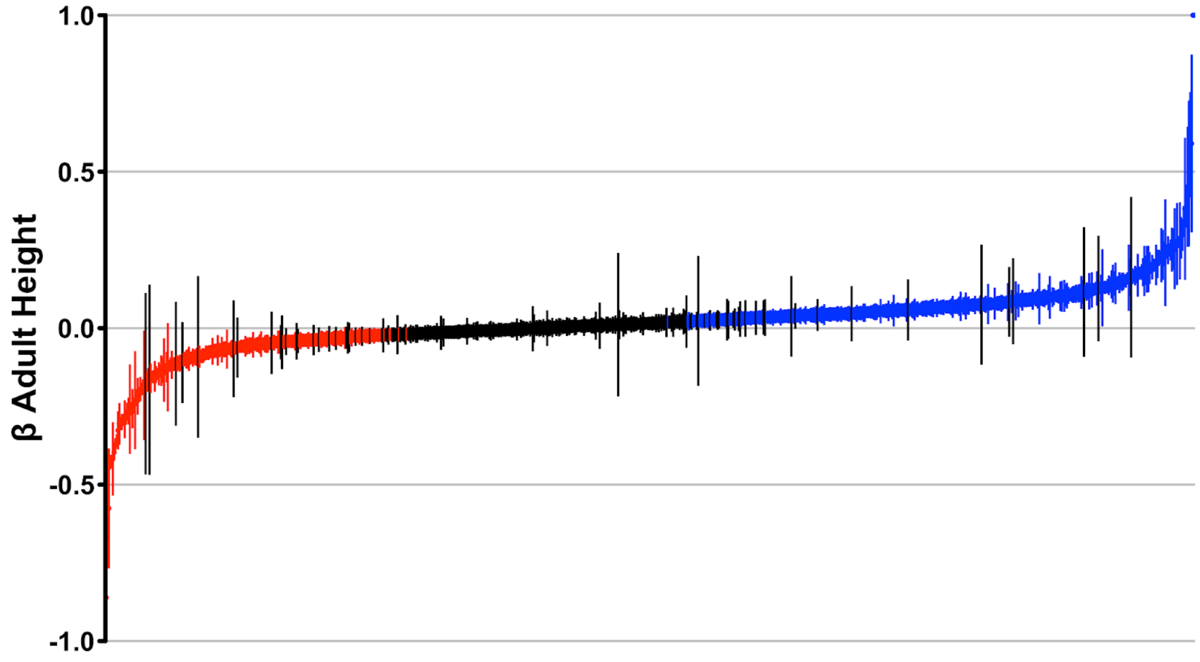


**Heterogeneity with height of SNPs associated with IGF-1.** Shown are Mendelian randomisation results for 784 independent genetic signals with a known association with circulating IGF-1 levels. SNPs are ordered on the x-axis by their β value for adult height. SNPs in red have a significantly negative association with adult height while SNPs in blue have a significantly positive association with adult height (y-axis). Note that the y-axis has been capped at -1 and +1; two SNPs above this cap in the upper-right hand corner have β's of 1.6 [1.3-1.8] and 1.9 [1.6-2.2]. Error bars represent the 95% confidence interval.
